# Low vision practice and service provision among Optometrists in Ghana: a nationwide survey

**DOI:** 10.1101/2023.07.16.23292720

**Authors:** Kwadwo Owusu Akuffo, Isaiah Osei Duah Junior, Eldrick Adu Acquah, Elna Abadua Mensa, Albert Kwadjo Amoah Andoh, David Ben Kumah, Bridget Senya Boateng, Josephine Ampomah Boateng, Kofi Osei-Poku, Werner Eisenbarth

**Author notes:** Correspondence Dr. Kwadwo Owusu Akuffo; Department of Optometry and Visual Science, Kwame Nkrumah University of Science and Technology, Kumasi, Ghana; or.

## Abstract

**Aim:** To characterize practice patterns of low vision services among Optometrists in Ghana.

**Methods:** The nationwide cross-sectional survey identified entities through the Ghana Optometrists Association (GOA) registry and utilized a semi-structured questionnaire to consolidate survey information that comprises practitioners’ demographics, available services, diagnostic equipment, barriers to service provision and utilization, and interventions.

**Results:** 300 Optometrists were identified, with 213 surveyed (71% response rate). About fifty percent (52.6%) were in private practice, and more than two-thirds (77%) did not provide low vision services. Most (≥ 70%) established lack of assistive devices, and basic eye care examination kits as the main barriers to low vision service provision. Similarly, practitioners reported unawareness of the presence of low vision centres (76.1%), and high cost of low vision aids (75.1%) as the prime perceived barriers for patients to utilize low vision services. Continuous professional development and public education (89-90%) were suggestive interventions to improve the uptake of low vision services. After statistical adjustment, private facility type (Adjusted odds ratio [AOR] = 0.35, p = 0.010) and lack of basic eye examination kits (AOR = 0.32, p = 0.002) were significantly associated with reduced odds of low vision service provision. Conversely, 15-19 years of work experience (AOR = 8.49, p = 0.022) were significantly associated with increased odds of low vision service provision.

**Conclusion:** Overall, the results indicate inadequate low vision coverage and service delivery. Government policies must be directed towards equipping practitioners with equipment and subsidize patient cost of treatment to optimize low vision care.

**What is already known on this topic?:** Fewer past studies have reported poor low vision service delivery in selected regions and hospitals in Ghana. However, this evidence is an under-representation of the coverage of low vision service delivery in the country and warrants a more robust design to obtain comprehensive estimates.

**What this study adds:** The current study extends the existing literature by providing extensive evidence on the practice pattern of low vision services, barriers, and interventions in Ghana.

**How this study might affect research, practice, or policy?:** The unmet needs of low vision service delivery for residual vision necessitate institutionalizing pragmatic strategies to augment low vision service delivery, uptake, and delivery in the region.

**Synopsis/Precis:** This paper highlights the scope of low vision practice in Ghana. The findings show an unmet low vision coverage, significantly influenced by practice settings, years of work experience, and rudimentary eye examination equipment.

## INTRODUCTION

The increasing burden of eye diseases of refractive and non-communicable origin in Ghana poses a remarkable national health concern owing to the adverse sequelae on the quality of life and productivity of afflicted patients [1 2]. Such vision deterioration often results in residual and/or low vision; defined by the World Health Organisation as visual acuity worse than 6/18 to light perception with optimum treatment or a visual field under 10 degrees from the fixation point. Unlike patients with optimal vision, visually impaired and/or low vision patients experience aberrant visual-motor coordination, that eventually result in difficulty in performing everyday activities, mobility and/or transportation challenges, and increased physical dependency [3 4]. These patients are at risk of a broad spectrum of psychological distress ranging from depression and anxiety that could potentially accentuate into suicide and even death [5].

Contrary to advanced countries, visually impaired and/or low vision patients in resource-constraint environments are predisposed to a vast unmet visual need. These trends are in part due to decreased doctor-to-patient distribution, inadequate eyecare facilities and medical resources for care givers, as well as the relatively high subsidies on medical treatments [6-8]. The prevalence of vision impairment and blindness in Ghana is presently on a rise, yet there remains inadequate low vision treatment and rehabilitation services to address such challenges [9]. Despite the markedly increased motivation of optometry students towards clinical practice, these trained eye care professionals are usually limited by the essential equipment to provide adequate primary eye care services including low vision care [8 10 11]. Although patients with sub-optimal vision usually resort to low vision care, the majority face challenges to access care [8]. Previous work by Kyeremeh and Mashige [8] provides baseline evidence on the scope of low vision services in Ghana. However, this data is hampered by its under-representation, given its exclusive enrolment of optometrists from only two of the hitherto ten; currently sixteen administrative regions of Ghana.

To extend the prevailing evidence, this nationally representative survey comprehensively describes the scope of practice and coverage of low vision services rendered by primary eye care providers in Ghana. Specifically, the study characterizes the practice patterns of low vision services among optometrists in Ghana. Overall, the study presents the state of low vision service delivery by optometric clinicians for clientele with residual vision in Ghana. The results provide substantive evidence for resource allocation, formulation, and implementation of strategic policies to improve low vision coverage.

## MATERIALS AND METHODS

The study was approved by the Committee on Human Research, Publication and Ethics of the Kwame Nkrumah University of Science and Technology, Kumasi, Ghana (CHRPE/AP/286/22) after formal permission was obtained from the leadership of the Ghana Optometric Association. Participants informed consent was obtained and all protocols were consistent with the tenets of the Declaration of Helsinki. The nationwide cross-sectional survey, conducted between January 2022 and September 2022, assessed the practice patterns of low vision services among optometrists in Ghana. As a lower-and-middle income country (a country with a total economic value between $1006 and $3955), Ghana has an estimated population of thirty-two million distributed across sixteen administrative regions [12 13]. The survey population constituted all optometrists registered under the Ghana Optometry Association (GOA), a nationally recognized professional regulatory body that oversees the practice and advancement of optometry in Ghana and integrates the activities of optometrists with the Allied Health Professions Council of Ghana (AHPC).

The structured questionnaire used for this survey was adapted from Kyeremeh and Mashige [8] and constituted both open and closed-ended questions which were either in electronic or hard copy versions. As appropriate, the questionnaire was administered to all participants either face-to-face or electronically via google forms [14]. The data collection form had questions aimed at identifying optometrists’ demographics, mode and scope of practice, standards of diagnosis, tests and procedures, treatment/management options, barriers, and a comments section for interventions to augment the delivery and uptake of low vision services.

### Statistical analysis

Statistical analyses were performed with Statistical Product and Service Solution (IBM Corporation IBM® SPSS® Statistics for Windows, Version 25.0 Armonk, NY) compatible with Windows 10. Descriptive statistics (frequencies and percentages) were used to summarize the characteristics of low vision services (demographics, services provided, equipment available, barriers and interventions). Association between predictor and outcome variables were explored using bivariate logistic regression, and all variables significant at p < 0.05 were selected for inclusion into the multivariate logistic regression model. Statistical significance was set at a p≤ 0.05 and at a 95% confidence interval.

## RESULTS

Out of the 300 eligible optometrists, 213 completed the survey (response rate: 71.0%). Most were males (63.0%), aged 30-39 years (53.1%), worked in private facility (52.6%) and with 1-4 years’ work experience (43.6%). The predominant barrier for not providing low vision services was lack of low vision assistive devices (77.5%) and lack of basic examination kit (71.4%). Suggestive interventions to optimize low vision care were continuous professional development (90.1%), specialized training on low vision care (90.1%), public education on low vision and low vision services (89.7%), making low vision assistive devices affordable (88.7%) and provision of low vision equipment for assessment (87.3%) as shown in Table 1.

**Table 1:** Description of the sample.

| Variable | N | % |
| --- | --- | --- |
| *Demographic characteristics* |  |  |
| Sex<sup>†</sup> |  |  |
| Male | 133 | 63.0 |
| Female | 78 | 37.0 |
| Age years<sup>†</sup> |  |  |
| 20-29 | 71 | 33.6 |
| 30-39 | 112 | 53.1 |
| 40-49 | 24 | 11.4 |
| 50-59 | 4 | 1.9 |
| Facility type |  |  |
| Public | 62 | 29.1 |
| Private | 112 | 52.6 |
| NGO | 5 | 2.3 |
| CHAG | 34 | 16 |
| Work experience<sup>†</sup> |  |  |
| 1-4 | 92 | 43.6 |
| 5-9 | 43 | 20.4 |
| 10-14 | 65 | 30.8 |
| 15-19 | 7 | 3.3 |
| 20-24 | 2 | 0.9 |
| 25 and above years | 2 | 0.9 |
| *Barriers of low vision service delivery* |  |  |
| Lack of personnel |  |  |
| Yes | 71 | 33.3 |
| No | 142 | 66.7 |
| Low profit from low vision care |  |  |
| Yes | 55 | 25.8 |
| No | 158 | 74.2 |
| Lack of basic eye examination kits |  |  |
| Yes | 152 | 71.4 |
| No | 61 | 28.6 |
| Lack of referral centres |  |  |
| Yes | 85 | 39.9 |
| No | 128 | 60.1 |
| Lack of low vision assistive devices |  |  |
| Yes | 165 | 77.5 |
| No | 48 | 22.5 |
| Labour Intensive |  |  |
| Yes | 67 | 31.5 |
| No | 146 | 68.5 |
| *Interventions to low vision service delivery* |  |  |
| Continuous professional development training |  |  |
| Yes | 192 | 90.1 |
| No | 21 | 9.9 |
| Specialized training on low vision care |  |  |
| Yes | 192 | 90.1 |
| No | 21 | 9.9 |
| Public education on low vision and low vision service delivery |  |  |
| Yes | 191 | 89.7 |
| No | 22 | 10.3 |
| Making low vision assistive devices affordable |  |  |
| Yes | 189 | 88.7 |
| No | 24 | 11.3 |
| Provision of low vision equipment for assessment |  |  |
| Yes | 186 | 87.3 |
| No | 27 | 12.7 |

‡, N ≠ 213 for all variables as some participants failed to respond to all items on the questionnaire; %, percentage frequency.

### Low vision services

The majority of Optometrists (79.8%) had patients that sought for low-vision care. However, only 23.0% provided low vision care which was limited to dispensing, training on use of assistive devices, and low vision rehabilitation (Table 2).

**Table 2:**
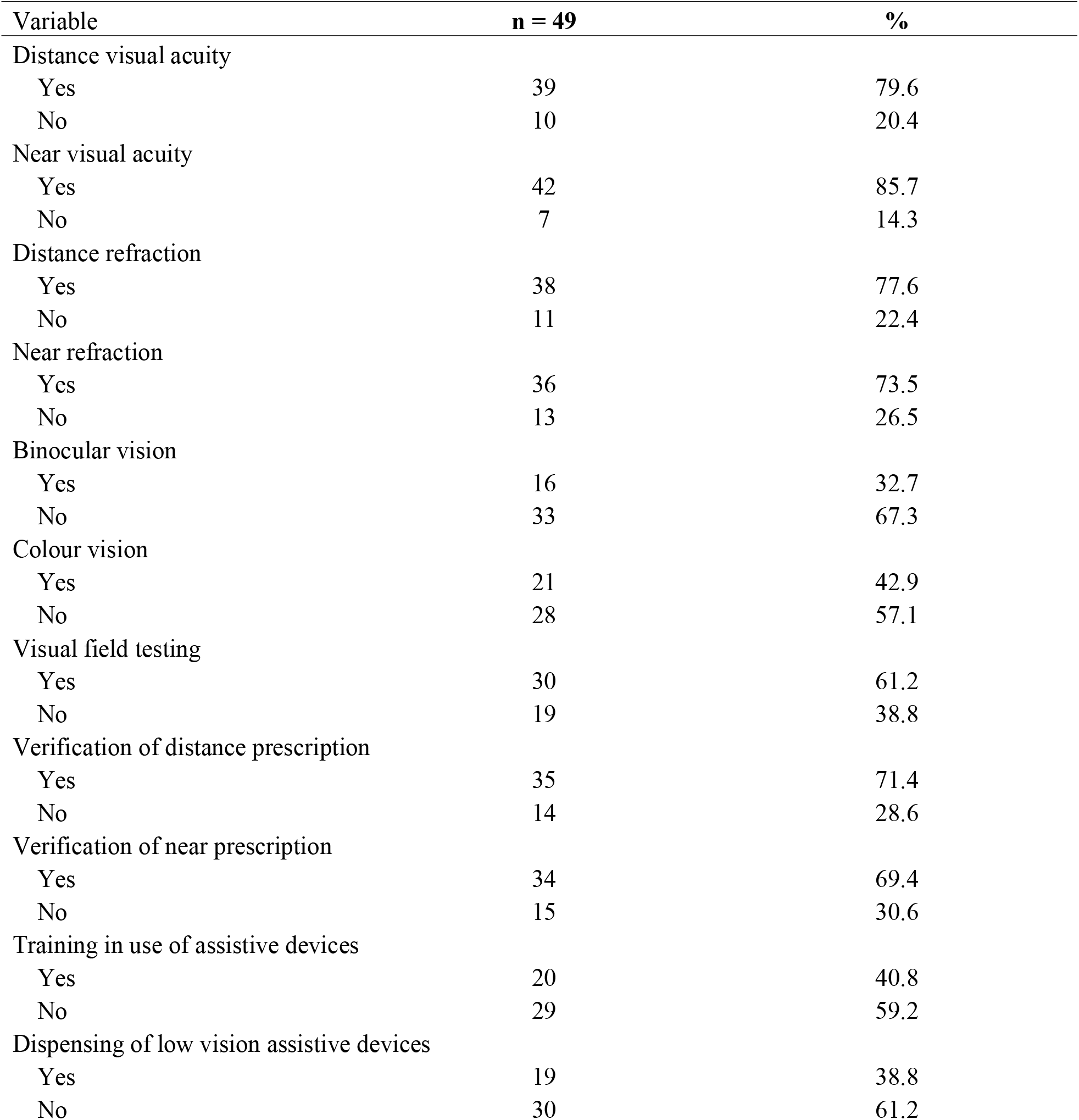
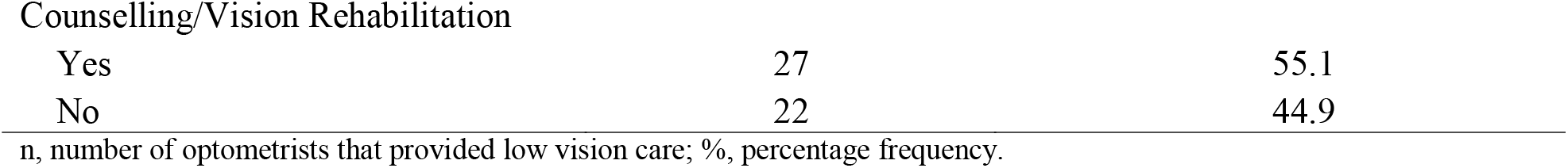
Low vision services.

| Variable | n = 49 | % |
| --- | --- | --- |
| Distance visual acuity |  |  |
| Yes | 39 | 79.6 |
| No | 10 | 20.4 |
| Near visual acuity |  |  |
| Yes | 42 | 85.7 |
| No | 7 | 14.3 |
| Distance refraction |  |  |
| Yes | 38 | 77.6 |
| No | 11 | 22.4 |
| Near refraction |  |  |
| Yes | 36 | 73.5 |
| No | 13 | 26.5 |
| Binocular vision |  |  |
| Yes | 16 | 32.7 |
| No | 33 | 67.3 |
| Colour vision |  |  |
| Yes | 21 | 42.9 |
| No | 28 | 57.1 |
| Visual field testing |  |  |
| Yes | 30 | 61.2 |
| No | 19 | 38.8 |
| Verification of distance prescription |  |  |
| Yes | 35 | 71.4 |
| No | 14 | 28.6 |
| Verification of near prescription |  |  |
| Yes | 34 | 69.4 |
| No | 15 | 30.6 |
| Training in use of assistive devices |  |  |
| Yes | 20 | 40.8 |
| No | 29 | 59.2 |
| Dispensing of low vision assistive devices |  |  |
| Yes | 19 | 38.8 |
| No | 30 | 61.2 |
| Counselling/Vision Rehabilitation |  |  |
| Yes | 27 | 55.1 |
| No | 22 | 44.9 |

n, number of optometrists that provided low vision care; %, percentage frequency.

### Low vision equipment and accessories

The majority of optometrists (30-94%) lacked low vision equipment and accessories for vision examination (see Figure 1). Most lacked precision vision (PV)-16 colour vision test (93.9%), hand disc perimeter (91.8%), tangent screen (81.6%), preferential looking system (79.6%) and light box for VA test (73.5%). Low vision optical and/or non-optical aids were limited in most optometric practices (see Figure 2).

### Barriers to utilisation of low vision services

The perceived barriers that prevent patients from utilizing low vision services included: unawareness of the presence of low vision centres (76.1%), high cost of low vision services (75.1%), socially unacceptable assistive devices (43.2%), and patients not seeing the need (35.2%).

### Factors associated with low vision services among Optometrist in Ghana

The bivariate logistic regression analysis showed that private facility type (compared with public facility type: odds ratios [OR] = 0.37, p = 0.007), 15-19 years’ work experience (compared with 1-4 years’ work experience: OR = 11.02, p = 0.006), and lack of basic eye examination kits (OR = 0.34, p = 0.002) were significantly associated with low vision services. In the multiple logistic regression analysis, private facility type (AOR = 0.35, p = 0.010 compared with public facility type) and lack of basic eye examination kits (AOR = 0.32, p = 0.002) were significantly associated with reduced odds of low vision services whereas 15-19years’ of work experience (compared with 1-4years’ of work experience: AOR = 8.49, p = 0.022) were significantly associated with increased odds of low vision services (Table 3).

**Table 3:**
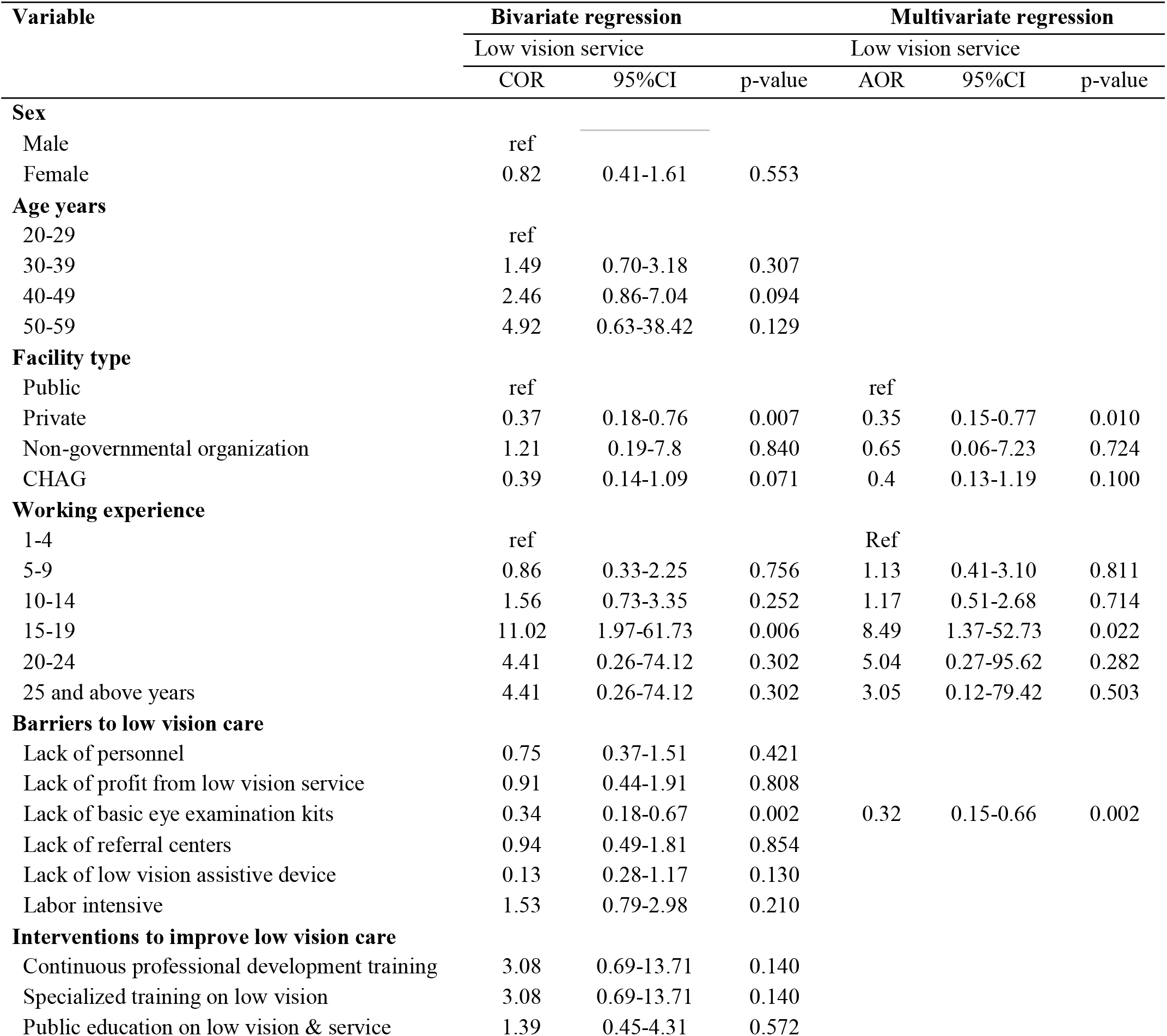
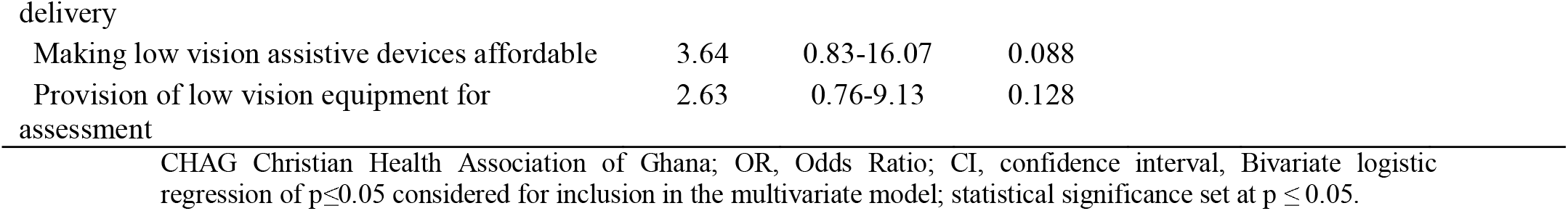
Bivariate and multiple regression of factors associated with low vision service.

## DISCUSSION

The survey reports on the practice patterns of low vision services among optometrists in Ghana and provide evidence on the practice setting, clientele services, equipment, vision aids, barriers and interventions to low vision services in Ghana. After statistical adjustment, private facility type, and lack of basic eye examination kits were significantly associated with reduced likelihood of low vision service provision whereas 15-19 years of work experience were significantly associated with increased likelihood of low vision service provision.

Optimal vision remains essential for performing daily activities (such as mobility, recognizing currency, and reading inscriptions on medicines) especially among students, working class, and geriatric population. Whereas efficient vision results in proficient productivity, uncorrected vision impairment culminate in physical dependency. Unlike elderly, childhood vision loss is worrying owing to the long years lived with disability and consequences on their career aspiration [15 16]. Residual and/or low vision presents a myriad of psychological consequences, notably shock, anxiety, denial, depression, withdrawal, and emotional acceptance. Consequentially, such visual handicap extends to negatively impact social interactions and further places an economic burden on self and family due to the cost of treatment and securing optical aids. The availability of accessible and proficient low vision treatment and rehabilitation support systems remains paramount and underscores optimizing residual vision to improve patient’s overall quality of life [17 18]. In addition, establishing appropriate referral pathways could improve referral for low vision treatment and rehabilitation services during the end stage of ocular lesions and putatively mitigate functional impairments and the associated mental distress [19-21]. However, the scope of practice of such services in Ghana is poorly understood. Therefore, the present study describes the practice patterns of low vision service delivery in Ghana.

About half (52%) of the practitioners that provide low vision care were in private optometric facilities. Specialised eye care services are generally skewed toward private facilities in Ghana [7 10 22]. For instance, previous work on the practice patterns of amblyopia by Acheampong et al. identified slightly less than half of optometrists that manage amblyopia were in private practice [10]. Similarly, a report by Mensah-Debrah and colleagues revealed that private facilities constitute fifty percent of the centres offering diabetic retinopathy treatment services [7]. Contrary to public facilities, private practices are well resourced with advanced arsenal for treatment/management and highly trained personnel for specialized care services [23 24]. However, the observed decreased likelihood of private facilities to provide low vision care might be attributed to the limited coverage of the National Health Insurance Scheme and the relatively high cost treatment of private specialized treatment services that could limit access to care[25].

More than two-thirds (77%) of practitioners did not provide low vision care service for clientele that sought for care. The results are comparable to a previous study by Akuffo et al. where only 23.8% of optometrists provided low vision care [22]. Although the access to basic ophthalmic care such as clinical examination and management, refraction, and optical dispensing were markedly higher than the present study, the findings here are not surprising as most existing clinical infrastructure are not adapted for specialized care including low vision [22]. Further, the limited low vision rehabilitation and counselling services remains a challenge, as uncorrected visual impairment could culminate in psychological distress [26].

Providing adequate low vision care remains paramount to preventing further functional impairment of visual function. However, most optometrists were found to lack basic eye examination kits, and this was associated with decreased likelihood of low vision care. The findings is consistent with an earlier work by Kyeremeh and Mashige, where most optometrists lacked equipment for low vision services [8]. Given the equipment constraints, most underlying pathologies causing residual vision could aggravate into complete blindness [27 28]. However, the observed increase likelihood of low vision services provision with 15-19 years of work experiences demonstrates the crucial role of experience practioners compensating for the equipment gap through their systematic approach to treatments.

The study found the availability of optical aids for patients with residual vision to be low, which is a perturbing situation. Despite substantial vision deterioration and functional impairment in low-vision patients, providing visual and non-optical assistive devices to optimize residual vision could reconcile and improve the quality of life of these patients [29].

The reported lack of awareness of low vision centres and the high purchasing cost of low vision assistive devices are consistent with previously published reports [30-32]. In mitigating these barriers, public education campaigns on low vision care and low vision centres, as well as increasing affordability of low vision aids, were suggestive interventions to improve the uptake of low vision services in this population.

## Strengths and limitation

Although the current work presents comprehensive evidence on the practice patterns of low vision services in Ghana, it is subjected to some few limitations. The report on the available resources for low vision treatment may be underestimated due to recall bias. Similarly, given the design of the study, intricate patterns on the service provisions were not explored in depth, however, the comprehensive nature of the data collection elicited essential parameters on the present scope and practice of low vision care services in Ghana.

## Conclusion

There is an unmet eye care need for patients with low vision in Ghana. The equipment to provide both basic and comprehensive low vision assessment is lacking leaving a considerable number of low vision clientele underserved. Given the prevailing evidence, we implore government, policy makers, corporate stakeholders of health to implement pragmatic strategies to equip optometrists with the requisite armamentarium for low vision care as well as subsidizing the cost of low vision assistive devices. Furthermore, the findings suggest the initialisation of health education and health promotion to improve low vision awareness. Future research programs should explore low vision rehabilitation and referral pathways and their potential enablers and barriers.

## Supporting information

Fig1_Low vision equipment and aaccessories.jpg

Fig 2_ Low vision optical and non-optical aids.jpg

## Contributions

KOA, WE., IODJ- conceptualization of the study; KOA, EAM, IODJ, EAA -– project administration; EAM and JAB, - data collection; IODJ, AKAA, and EAM - statistical analysis and data visualization; KOA, IODJ, EAA, BSB, AKAA, DBK, BSB, JAB, KOP and WE; data interpretation; KOA, IODJ, EAA, BSB and WE- drafting original manuscript; KOA, IODJ, EAA, BSB, AKAA, DBK, BSB, JAB, KOP and WE- review for important intellectual merits; KOA and WE- supervision; KOA and WE correspondence.

## Patient and public involvement

Patients and/or the public were not involved in the design, conduct, reporting, or dissemination plans of this research.

## Patient Consent

All participants agreed and gave their consent to partake in the study after the aims and potential benefits and risks were well spelt out.

## Ethics approval

The study was conducted according to the Tenets of the Declaration of Helsinki and ethical approval was obtained from the Committee on Human Research, Publication and Ethics, Kwame Nkrumah University of Science and Technology, Kumasi, Ghana (CHRPE/AP/286/22). Approval was also obtained from the president of the Ghana Optometric Association.

## Data availability statement

The datasets used and analysed during the current study are available from the corresponding authors upon reasonable request.

