## Supplementary figures and images for "Low vision practice and service provision among Optometrists in Ghana: a nationwide survey"

### Fig1_Low vision equipment and aaccessories.jpg

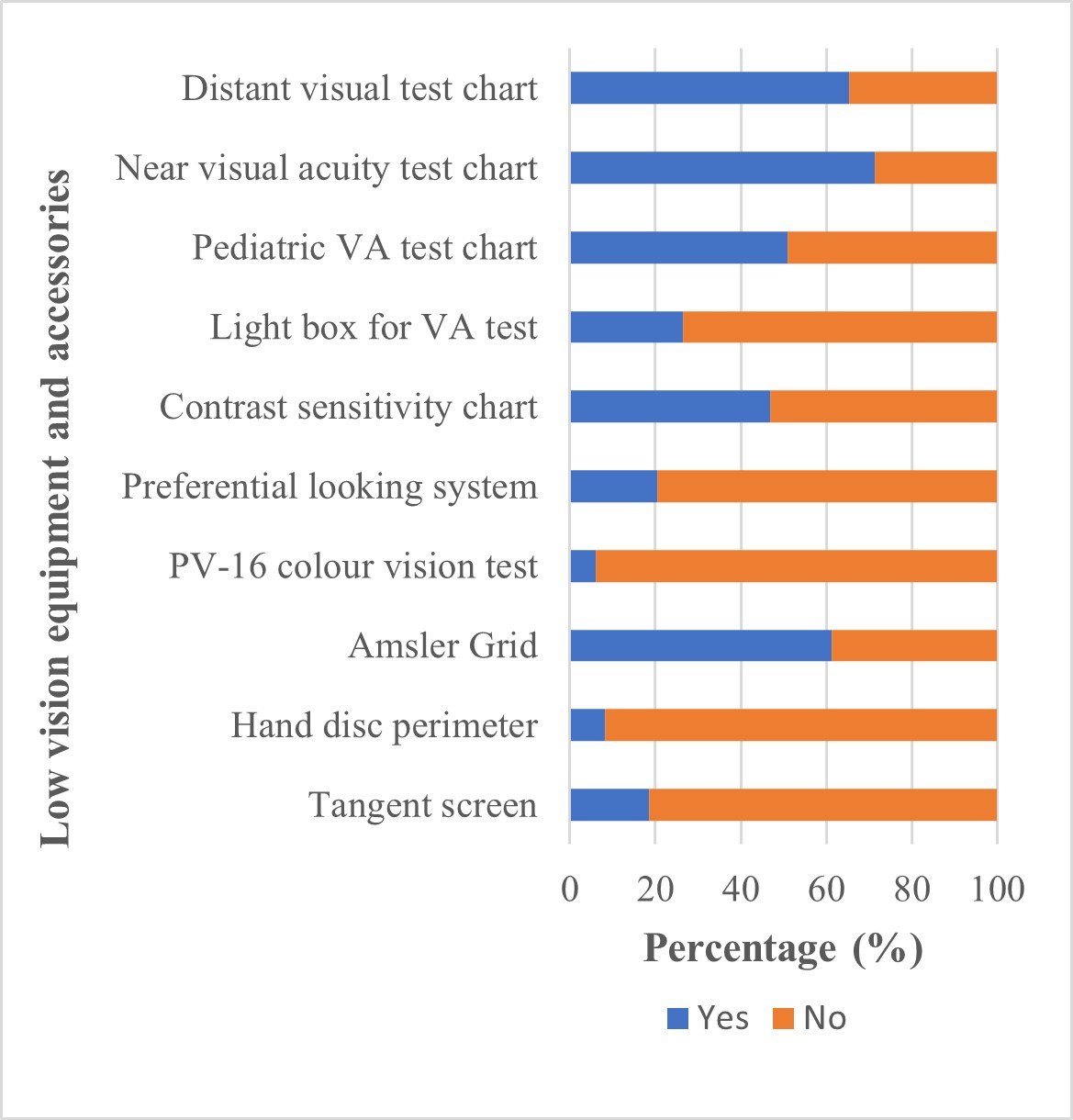

### Fig 2_ Low vision optical and non-optical aids.jpg

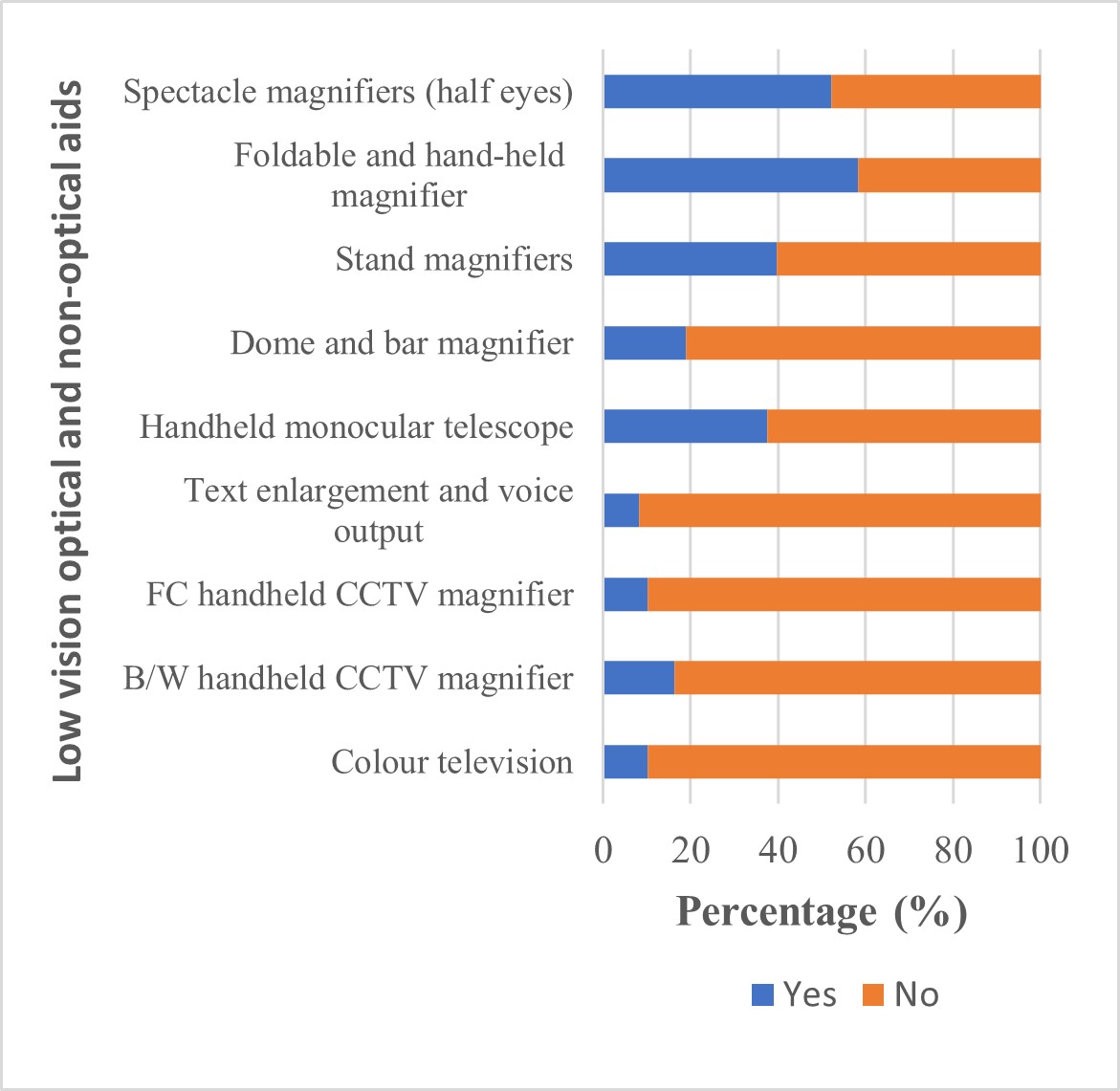
